# Predicting COVID-19 case status from self-reported symptoms and behaviors using data from a massive online survey

**DOI:** 10.1101/2023.02.03.23285405

**Authors:** Mashrin Srivastava, Alex Reinhart, Robin Mejia

**Affiliations:** Machine Learning Department, Carnegie Mellon University; Department of Statistics & Data Science, Carnegie Mellon University

## Abstract

With the varying availability of RT-PCR testing for COVID-19 across time and location, there is a need for alternative methods of predicting COVID-19 case status. In this study, multiple machine learning (ML) models were trained and assessed for their ability to accurately predict the COVID-19 case status using US COVID-19 Trends and Impact Survey (CTIS) data. The CTIS includes information on testing, symptoms, demographics, behaviors, and vaccination status. The best performing model was XGBoost, which achieved an F1 score of *≈* 94% in predicting whether an individual was COVID-19 positive or negative. This is a notable improvement on existing models for predicting COVID-19 case status and demonstrates the potential for ML methods to provide policy-relevant estimates.

## 1 Introduction

The COVID-19 pandemic has affected over 675.6 million people worldwide and caused over 6.76 million deaths (Worldometers, 2023), as of 2 Feb 2023. In the United States alone, there have been 104.3 million confirmed cases and 1.13 million deaths, but the actual number of cases is expected to be significantly higher, as confirmed cases only include those with a positive test, and tests have not been universally available throughout the pandemic. RT-PCR testing is the standard procedure for diagnosing COVID-19, but it requires specialized materials and personnel, which may not always be readily available. The use of home-based antigen tests, which are not required to be reported to public health agencies, has also contributed to a decline in the utilization of RT-PCR tests.

Multiple studies have found that ML algorithms can be used to screen for COVID-19 using x-ray and CT scans (Zargari et al., 2021; Ismael & Şengür, 2021; Panwar et al., 2020; Hussain et al., 2021; Roberts et al., 2021; He et al., 2020; Afshar et al., 2021; Song et al., 2021). However, such data is difficult to obtain (Zhao et al., 2020; Shakouri et al., 2021), as an estimated two-thirds of the world lacks access to it (Mbewe et al., 2020). In Brinati et al. (2020) and Ferrari et al. (2020), routine blood test data was used for predicting COVID-19. The limitations of these studies include a small number of patients (160–300 samples), and data obtained from a single center (Kukar et al., 2021).

Predictive models based on symptoms can possibly improve the understanding of COVID-19 trends, as official case counts based on testing are likely to be an undercount, and the level of reporting may vary over time and space, depending on pandemic conditions. Collecting symptom data is also relatively simple and inexpensive. However, the multitude of symptoms related to COVID-19, many of which resemble other common viruses, poses a difficulty in determining which symptoms are most indicative of COVID-19 infection. The use of machine learning techniques allows for the recognition of symptom patterns that are suggestive of COVID-19. Zoabi et al. (2021) employed a gradient-boosting model using decision-tree base-learners to predict COVID-19 infection based on symptoms, age, sex, and known contact with infected individuals, attaining 87.30% sensitivity and 71.98% specificity. Gomathi et al. (2020) used similar features, along with details regarding international travel to predict COVID-19 in India. Maharaj et al. (2021) (in Canada) and Menni et al. (2020) (in the UK) also used symptoms to predict COVID-19. One limitation of these studies is the use of small datasets (100–100,000 samples). In Rufino et al. (2022), the authors used a large survey like the one we will present. A random forest classifier was trained for multiple countries and performed at 31% to 71% F1 score, depending on the country.

## 2 Methods

### 2.1 Data Acquisition And ParticipantS

The data used in this study is from the US COVID-19 Trends and Impact Survey (CTIS), a large, cross-sectional, internet-based survey (Salomon et al., 2021). CTIS respondents were Facebook users aged 18 and older who were randomly sampled from the Facebook Active User Base and invited to complete the survey, which was administered by Carnegie Mellon University. CTIS collected information including COVID-19 symptoms, risks, behaviors, testing and vaccination. The large scale of the survey ensures that the sample captures a large number of individuals who have taken COVID-19 tests and allows training and evaluating the model on a large sample. For this study, we used the data from May 2021 to February 2022, which includes 12.41 million responses.

### 2.2 Data Pre-processing AND Labeling

Because of the limitations of large-scale online surveys, we attempted to identify and remove responses that showed potential “trolling” behavior. Trolling refers to respondents who did not answer the survey questions thoughtfully or with genuine intent. Some of the methods we used to identify and remove these responses included identifying respondents who speed through the survey, select the same answer repeatedly, choose answers to create a specific pattern, select all options for multiple choice questions, provide unrealistic or inconsistent answers, or provide nonsensical inputs to text response questions. Approximately 4.8% of responses were identified as showing potential trolling behavior and were removed from the study.

The ground truth label was a binary label categorizing a respondent as positive or negative, based on self-report data on the results of a test for COVID-19. A positive label was assigned if the respondent had both tested for COVID-19 in the last 14 days and tested positive in their most recent COVID-19 test. If the respondent tested tested negative for COVID-19 in the last 14 days, a negative label was assigned. The respondents who did not take a COVID-19 test in the last 14 days were not considered for this study.

### 2.3 Feature Selection

To evaluate the performance of our models, we divided the data into train, validation, and test sets in a 70:15:15 ratio. Moreover, each response to the survey includes many questions designed to meet a large number of policy and prediction needs.^1^ We used XGBoost to determine the most discriminative features to be used in conjunction with symptoms for predicting COVID-19 case status. The importance of each feature was determined by averaging the feature’s importances over all trees and weighting the frequency of feature usage in tree splits by the square of the improvement in model performance. The features we extracted for this study were testing reason, behavior features, and symptoms. Testing reason was determined by the respondent’s reason for taking a COVID-19 test, recorded as a categorical variable with three possible values: symptomatic, asymptomatic (e.g. they were required to be tested by an employer and had no symptoms), and contact with a COVID-19 patient. The remaining features were binary indicators for the presence or absence of various symptoms, as listed in the Appendix. We also evaluated the impact of including demographics and vaccination status on model performance, to compare with existing research.

### 2.4 Model Configuration

Our baseline model was based on Rufino et al. (2022), who used data from the international version of the COVID Trends & Impact Survey. They trained a random forest classifier using features related symptoms, newness of the symptoms, testing reason, vaccination status, behavior, occupation, demographics, area type. However, our study did not have access to certain features that were used in the baseline model, such as area type, number of people sleeping in the same place, and number of rooms at the place of residence, as these were not asked in the US version of the survey. Therefore, we did not include these features in our model.

We created five different configurations using the features we identified in Section 2.3: 1. Symptoms alone; 2. Symptoms and testing reason (Symptomatic, Asymptomatic, Contact); 3. Symptoms, testing reason, recent behaviors (masking, social distancing, out of state travel); 4. Symptoms and demographics (gender, pregnancy, age, highest degree of education, race); 5. Symptoms and vaccination status (yes/no to at least one vaccination).

To address the issue of class imbalance (more negative cases than positive cases) in the training data, the SMOTE algorithm (Chawla et al., 2002) was applied to generate synthetic samples of the minority class. To compare the performance of different machine learning algorithms, we used different models including AdaBoost, gradient boosting, logistic regression, multi-layer perceptron (MLP), decision trees, Gaussian Naive Bayes, quadratic discriminant analysis (QDA), random forests, XGBoost and a custom interpretable model. For this interpretable model, we trained an XGBoost model using the symptoms as the features and calculated the feature importance for each symptom. The feature importance scores were then re-scaled to a small number and used as weights for each symptom in the model. Extensive hyperparameter tuning was conducted using grid search with stratified 5-fold cross-validation, which is described in the appendix.

## 3 Result and Discussion

For the baseline model based on Rufino et al. (2022) using a random forest classifier with the features described above, we achieved an F1 score of 52.

The performance of the models for different modeling configurations mentioned in Section 2.4 is shown in Table 1. Note that accuracy scores are generally higher than F1 scores because we have imbalanced classes. Additionally, we found that using SMOTE did not improve model performance, so we present the results without using SMOTE.

**Table 1:** Performance for various models for different modelling configuration

| Model | Symptoms |  | Symptoms + Test Reason |  | Symptoms + TR + Behavior |  |
| --- | --- | --- | --- | --- | --- | --- |
|  | Accuracy | F1-Score | Accuracy | F1-Score | Accuracy | F1-Score |
| AdaBoost | 84.7 | 53.2 | 84.8 | 53.3 | 89.2 | 71.8 |
| GradientBoosting | 72.4 | 55.7 | 72.7 | 55.9 | 89.9 | 74.8 |
| LogisticRegression | 73.7 | 53.7 | 73.4 | 53.6 | 87.2 | 65.3 |
| MLP | 84.9 | 56.1 | 85.6 | 56.4 | 90.0 | 75.0 |
| Decision Tree | 84.0 | 54.6 | 84.7 | 54.9 | 84.9 | 64.5 |
| Gaussian NB | 80.6 | 58.1 | 81.2 | 58.3 | 81.7 | 60.8 |
| QDA | 78.5 | 56.2 | 78.4 | 56.2 | 79.1 | 59.4 |
| Random Forest | 84.3 | 54.8 | 84.8 | 55.1 | 89.4 | 73.9 |
| XGBoost | **84.8** | **87.1** | **85.1** | **87.2** | **89.8** | **93.6** |

Our model performed better than the baseline in all cases except when we used only symptoms and vaccination status as features. In this specific scenario, the baseline model using a random forest classifier performed better than our random forest classifier. The reason for the reduced performance of our model in this case may be because the baseline model uses all relevant symptoms, testing reason, vaccination status, behavior, occupation, demographic, and area type information, while our model only considers symptoms and vaccination status. However, it is worth noting that even in this case, our best performing model (XGBoost) outperformed the baseline model.

Our experimental results showed that the XGBoost classifier consistently outperformed the other models in all configurations, with the most notable difference in the case when the behavior attributes were not included. The highest performance was achieved in the configuration when the combination of symptoms, testing reason, and behavior features was used. Interestingly, the testing reason feature did not significantly contribute to the overall performance of the model, indicating that our model is not biased towards any particular reason for taking a COVID-19 test and performs equally well for symptomatic, asymptomatic, and contact with COVID-19 patient cases. However, behavioral features such as masking, social distancing, and out-of-state travel significantly improved model performance. These results suggest that the inclusion of behavior information can improve the accuracy of our model for predicting COVID-19 status.

We also evaluated the impact of demographic attributes and vaccination status on model performance, as shown in Table 2. Our results indicated that demographic information did not significantly improve model performance and is therefore not considered to be an important feature for predicting COVID-19. Moreover, we found that including vaccination status resulted in a decrease in model performance, particularly in terms of F1 score. XGBoost was the only model that showed a slight improvement in F1 score when vaccination status was included, but it also built a more complex tree in this case. Our analysis revealed that while the inclusion of vaccination status improved the model’s ability to predict COVID-19 negative cases, it performed worse in predicting positive cases. Overall, our results suggest that vaccination status does not improve prediction of COVID-19 when used with other features.

**Table 2:** Evaluating the contribution of demographics and vaccination status features

| Model | Symptoms + Demographics |  | Symptoms + Vaccination |  |
| --- | --- | --- | --- | --- |
|  | Accuracy | F1-Score | Accuracy | F1-Score |
| AdaBoost | 84.8 | 53.4 | 82.3 | 45.7 |
| Gradient Boosting | 72.3 | 55.6 | 82.4 | 48.2 |
| Logistic Regression | 73.8 | 53.7 | 82.3 | 43.5 |
| MLP | 84.8 | 56.2 | 82.3 | 45.7 |
| Decision Tree | 83.9 | 54.5 | 82.4 | 48.2 |
| Gaussian NB | 80.6 | 58.2 | 82.3 | 48.1 |
| QDA | 78.4 | 56.1 | 82.1 | 47.7 |
| Random Forest | 84.3 | 54.7 | 82.6 | 48.1 |
| XGBoost | **84.9** | **87.3** | **82.5** | **89.4** |

We also conducted an experiment to evaluate the ability of our model to generalize to future data. Specifically, we used the XGBoost model trained with symptoms, testing reason, and behavior features on data from January 2021 to February 2022 (12.46 million records) to predict COVID-19 for respondents from March 2022 to June 2022 (1.94 million records). The F1 score for this model was 93.4%. Also, we trained a similar model using data from March 2022 to June 2022 and tested it on respondents from the same time period and respondents from 2021. The F1 score for this model was 93.7% and 92.5% for the respective test sets. These results demonstrate the robustness and generalizability of our model, as it performs well across different time periods. It’s important to note that the data was generated from the same survey; however, this suggests that our model could potentially be used to predict COVID-19 cases in the future.

Finally, for the interpretable model mentioned in Section 2.4, the motivation was to develop a chart like PHQ-9 (Kroenke et al., 2001). This can help the clinicians to assess COVID-19 result quickly and easily. Symptoms such as fever, cough, tiredness/exhaustion, loss of taste and smell, chills had a weight of 1; stuffy or runny nose and muscle or joint aches had a weight of 2; shortness of breath, difficulty breathing, sore throat, persistent pain or pressure in the chest and headache had a weight of 3; and nausea/vomiting and diarrhea had a weight of 4. If the overall added score was less than 10, the model predicted a negative COVID-19 result, or else, it predicted a positive result. The F1 score for this simple, interpretable model was 65%.

## 4 Conclusion

This study demonstrates symptoms, testing reason and behavior can be used to predict COVID-19. We compared various ML models and found that the XGBoost achieved the highest F1 score of 93.6% in predicting COVID-19. To the best of our knowledge, this model was trained on the largest available health survey data and demonstrated consistent performance even for periods it was not specifically trained for. The strong performance of our model gives us confidence in its ability to accurately predict COVID-19, and if applied on a larger scale, it could help improve understanding of trends in COVID-19 cases in the US. One of the limitations of this study, however, is that the sample of individuals tested for COVID-19 is not a random sample from the population of respondents. These models are also limited by the quality and availability of data on symptoms, which can vary over time and location. Further work is needed to determine how generalizable the model is to the full population.

## Data Availability

Survey microdata are not publicly available because survey participants only consented to public disclosure of aggregate data, and because the legal agreement with Facebook governing operation of the survey prohibits disclosure of microdata without confidentiality protections for respondents. Deidentified microdata are available to researchers under a Data Use Agreement that protects the confidentiality of respondents. Access can be requested online (https://cmu-delphi.github.io/delphi-epidata/symptom-survey/data-access.html). Requests are reviewed by the Carnegie Mellon University Office of Sponsored Programs and Facebook Data for Good. County and state-level aggregates of key variables are publicly available in the COVIDcast API, and are presented in an interactive online dashboard (https://delphi.cmu.edu/covidcast/survey-results/?date=20211103).

## A Appendix

### A.1 Ethics

The COVID-19 Trends & Impact Survey was approved by the Carnegie Mellon University (CMU) Institutional Review Board, under protocol STUDY2020 00000162. All respondents gave informed consent before participating in the survey. The data acquisition design safeguards respondent privacy by ensuring that researchers at CMU do not receive an identifying information about respondents and Meta (formerly Facebook) does not see survey microdata.

### A.2 List of Symptoms

Fever, cough, shortness of breath, difficulty breathing, tired/exhaustion, stuffy or runny nose, muscle or joint aches, sore throat, persistent pain or pressure in your chest, nausea or vomiting, diarrhea, loss of smell or taste, chills, and headache.

### A.3 Model Configuration: Full List of Questions

#### A.3.1 For the Baseline

For the baseline model, based on Rufino et al. (2022), the authors used the international version of the COVID-19 Trends and Impact Survey (CTIS) data. The full list of questions can be found here: https://gisumd.github.io/COVID-19-API-Documentation/docs/survey instruments.html.

#### A.3.2 For the proposed work

For training and evaluating the models in this study, we used the United States version of the COVID-19 Trends and Impact Survey (CTIS) data. The US CTIS was conducted by Carnegie Mellon University (CMU) in Collaboration with Meta (previously Facebook). The full list of questions for US CTIS can be found here: https://cmu-delphi.github.io/delphi-epidata/symptom-survey/coding.html

### A.4 Hyperparameters for the models

**1. Adaboost**: n estimators = 1000, learning rate = 0.05 and algorithm = SAMME.R.

**2. Gradient Boosting**: loss = log loss, learning rate = 0.01, n estimators = 1500.

**3. Logistic Regression**: penalty = L1, C = 0.01.

**4. Multi-Layer Perceptron (MLP)**: activation = relu, solver = adam, alpha = 0.0001.

**5. Decision Tree**: criteria = gini, splitter = best.

**6. Gaussian Naive Bayes**: Used default parameter values in scikit-learn (Buitinck et al., 2013).

**7. Quadratic Discriminant Analysis (QDA)**: Used default parameter values in scikit-learn.

**8. Random Forest**: criterion = gini, n estimators = 2000.

**9. XGBoost**: obj = reg:squarederror, booster = gbtree, gamma = 0, max depth = 4, lambda = 7.

### A.5 Evaluation metrics

Since our data set is imbalanced, with more COVID-19 negative cases than COVID-19 positive cases, we choose F1 score as the evaluation metric for this study. We use the F1 score as the evaluation metric for this study.

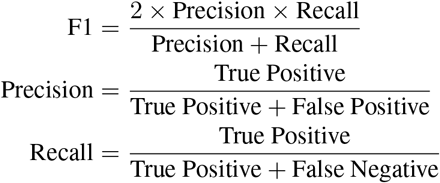

## Footnotes

1 Codebooks documenting all survey questions are available at https://cmu-delphi.github.io/delphi-epidata/symptom-survey/coding.html

